# Vaccine Confidence and Hesitancy at the start of COVID-19 vaccine deployment in the UK: An embedded mixed-methods study

**DOI:** 10.1101/2021.07.13.21260425

**Authors:** Chrissy h Roberts, Hannah Brindle, Nina T Rogers, Rosalind M Eggo, Luisa Enria, Shelley Lees

## Abstract

**Introduction:** Approval for the use of COVID-19 vaccines has been granted in a number of countries but there are concerns that vaccine uptake may be low amongst certain groups.

**Methods:** This study used a mixed methods approach based on online survey and an embedded quantitative/qualitative design to explore perceptions and attitudes that were associated with intention to either accept or refuse offers of vaccination in different demographic groups during the early stages of the UK’s mass COVID-19 vaccination programme (December 2020). Analysis used multivariate logistic regression, structural text modelling and anthropological assessments.

**Results:** Of 4,535 respondents, 85% (n=3,859) were willing to have a COVID-19 vaccine. The rapidity of vaccine development and uncertainties about safety were common reasons for COVID-19 vaccine hesitancy. There was no evidence for the widespread influence of mis-information, although broader vaccine hesitancy was associated with intentions to refuse COVID-19 vaccines (OR 20.60, 95% CI 14.20-30.30, p<0.001). Low levels of trust in the decision-making (OR 1.63, 95% CI 1.08, 2.48, p=0.021) and truthfulness (OR 8.76, 95% CI 4.15-19.90, p<0.001) of the UK government were independently associated with higher odds of refusing COVID-19 vaccines. Compared to political centrists, conservatives and liberals were respectively more (OR 2.05, 95%CI 1.51-2.80, p<0.001) and less (OR 0.30, 95% CI 0.22-0.41, p<0.001) likely to refuse offered vaccines. Those who were willing to be vaccinated cited both personal and public protection as reasons, with some alluding to having a sense of collective responsibility.

**Conclusion:** Dominant narratives of COVID-19 vaccine hesitancy are misconceived as primarily being driven by misinformation. Key indicators of UK vaccine acceptance include prior behaviours, transparency of the scientific process of vaccine development, mistrust in science and leadership and individual political views. Vaccine programmes should leverage the sense of altruism, citizenship and collective responsibility that motivated many participants to get vaccinated.

## INTRODUCTION

In December 2020, the United Kingdom became the first country to approve the use of a vaccine directed against the SARS-Cov-2 after successful trials of the Pfizer/BioNTech BNT162b2 mRNA vaccine [1]. Regulatory approval of the Oxford University/AstraZeneca ChAdOx1 [2] and Moderna mRNA-1273 [3] products brought two additional vaccines to the UK market by early January 2021. Immediately after the licensing of BNT162b2, the UK government commenced an ambitious national vaccination campaign that aimed to maximise the short term impacts by (1) leveraging the probability that there would be high levels of efficacy from a single dose of (any available) COVID-19 vaccine and (2) by delivering available vaccine to older and clinically vulnerable individuals first [4]. The UK’s gamble on the effectiveness of a single dose was vindicated by subsequent evidence that a single dose of BNT162b2 was highly protective against emergency hospitalisation and mortality, whilst a single dose of ChAdOx1 similarly protected from severe disease [5]. Early findings from passive surveillance of household transmission in England [6] also showed that vaccination was associated with a reduced secondary attack rate, suggesting that vaccinated people who contracted SARS-CoV-2 infections were less able to transmit infection than unvaccinated people [7]. The UK’s vaccination strategy and programme has been hailed a success, with three-quarters of all UK adults having received at least one dose of a SARS-CoV-2 vaccine and 50% having received two doses by 2021-03-17 [8]. This figure is somewhat higher than the estimates of a June 2020 survey of around 13,000 people in 19 countries, which reported that on average 71% of respondents were either likely or very likely to accept a SARS-CoV-2 vaccine. The very high uptake of SARS-CoV-2 vaccines in the UK appears to indicate a substantial increase in SARS-CoV-2 vaccine confidence and also seems to fly in the face of both (1) that these vaccines remain mostly uncharacterised for long-term safety and (2) that there has been a recent national and global trend of vaccine hesitancy for a range of vaccines [9] which has emerged as one of the most significant and complex public health challenges of the 21st century.

The WHO SAGE Working Group on Vaccine Hesitancy previously developed the “3 Cs” model of vaccine hesitancy [10] which describes three key factors [Confidence, Complacency, Convenience] that contribute to vaccine hesitancy. The success of the UK’s SARS-CoV-2 vaccination strategy can potentially be explained as having made the process of vaccination highly *convenient* by making a sufficient number of vaccine doses available through the National Health Service (NHS), at local health centres and at zero cost to the public. The level of *complacency* is also likely to be very low because of the highly visible personal, social, cultural, economic and global impacts of the pandemic. The drivers and extent of UK *confidence* in SARS-CoV-2 vaccines are however more debatable [11] and in this work we aim to understand how individuals living in the UK made decisions about their intentions to either accept or refuse vaccination at the very beginning of the UK national vaccination programme that commenced in December 2020. In order to achieve this goal we carried out an online survey of ∼4,500 adults living in the UK and applied an embedded mixed-methods approach to analysis and interpretation.

## METHODS

### Survey Design

We designed and deployed an embedded mixed-methods online survey as previously described [12]. Briefly, the survey included both quantitative and qualitative (open-ended text) questions that were relevant to the UK COVID-19 outbreak, COVID-19 vaccines and their relationship to participants’ health, health behaviours and attitudes. All data were collected anonymously and securely using ODK [13]. The survey was advertised using Facebook’s premium “Boost Post” feature [14] and ran from 2020-12-08 to 2020-12-16. All questions in the survey were voluntary, meaning that participants could skip questions that they did not wish to answer. Adverts were targeted to the eligible population of people aged 18 and over and living in the UK. All participants were asked to provide informed consent and to confirm their eligibility.

The survey included questions on the topics of (1) Demographics, (2) Compliance with testing and isolation following COVID-19 symptoms, (3) Use of the NHS COVID-19 contact tracing app, (4) Trust in the government and their decision making, (5) Health condition and exercise, (6) Domestic and gender-based violence and (7) Attitudes towards vaccination.

Study respondents were asked “When COVID-19 vaccines become available, will you be happy to get vaccinated?” [Yes | No], “Why would/wouldn’t you get vaccinated?” [Free text] and “Which of these best describes your general feelings about vaccines?” [I believe that vaccines are an effective way to control infectious diseases | I have concerns about the usefulness of some or all vaccines, but have not turned down the offer of vaccination for me or a member of my household | I have concerns about the usefulness of some or all vaccines and have previously turned down one or more offers of vaccination for me or a member of my household]. To gauge previous vaccine seeking behaviours, we asked participants “Do you get a seasonal flu vaccine?” [Always/Usually | Sometimes | Never]. Participants indicated if they were disabled as described in section 6 of the UK Equality Act 2010 [Yes | No] and provided a self-assessment of their general health over the last four weeks [Very bad | Bad | Fair | Good | Very good].

Respondents were further asked “Do you think that the UK government tells you the whole truth about coronavirus and COVID-19?” [Yes/No], “Do you think that the UK government tells you the whole truth about coronavirus and COVID-19” [Always | Mostly | Sometimes | Almost never | Never | I don’t know] and “Do you feel that your household has experienced financial hardship as a result of COVID-19” [Yes | No].

Demographic variables included in the analysis were age group [18 - 29 years, 30 - 49 years, 50 - 69 years and 70+], gender [Female | Male | Another gender], highest educational attainment [GCSEs/O-levels | A levels/Highers | Degree or higher degree], ethnicity [using UK Government 2011 census groups [15]] employment [Full time | Part time | Home-maker | Retired | Student | Unemployed], annual household income [Less than £15,000 | £15,000 - £24,999 | £25,000 - £39,999 | £40,000 - £59,999 | £60,000 - £99,999 | £100,000+], political views [Conservative/Right” | Floating/Centre | Liberal/Left] and UK postcode area. Participants also provided information on whether they had prior doctor-diagnoses of several medical conditions including asthma, cancer, type 1 and 2 diabetes, heart disease, hypertension, lung disease, obesity, stroke, depression and anxiety. To assess recent signs of acute depression, participants were asked “How often did you feel down, depressed, or hopeless in the past four weeks [Not at all | Less than half the time | More than half the time | Every day].

### Analysis

Analysis was performed using R version 4.0.3. Missing data were imputed using the R package ‘mice’ [16]. For downstream analysis, 15 imputed data sets were pooled using the R package ‘sjmisc’ to find the modal value for each missing data point. The number of survey responses from ethnic minority groups was low and for statistical purposes we categorised ethnicity into two classes [White | Ethnic minority]. Acute depression was classified as none [Not at all], mild [less than half the time] and moderate-severe [More than half the time | Every day]. Univariate binomial logistic regression was used to determine whether there was evidence for an association between each variable and the outcome of vaccine acceptance. Any variable that was significantly associated (after false discovery correction for multiple testing) with vaccine acceptance decisions in univariate analysis (Q < 0.05) was then included in a multivariate binomial logistic regression analysis.

### Topic modelling

Structural topic modelling (STM) of open-ended text was used to identify key topics associated with acceptance and non-acceptance of a COVID-19 vaccine using the R package ‘stm’ [17]. Free text responses to the question “Why would/wouldn’t you accept a COVID-19 vaccine?” were processed into separate text corpuses [accept/refuse] and analysed individually. Common stopwords (and, is, don’t, can’t etc.) and semantically neutral words such as “COVID” and “Vaccine” were removed. The number of topics that were included in each analysis was determined from visualisation of charts comparing several key indicators provided by the STM process, including semantic coherence (e.g. the clarity of topics), held-out likelihood (e.g. likelihood of retaining documents in the analysis) and minimal residuals of the model fit. When identifying the optimal number of topics to include in the analysis, we sought to maximise the held-out likelihood (number of documents retained) whilst minimising the residuals. STM assigned a set of scores (theta scores) to each free-text response or ‘quote’ in the corpus and these scores could be used to assign each quote to one or more of the topics. We extracted the highest scoring quotes (theta > 0.3) for each topic and used a lazy-consensus approach to defining the topic names that we felt best described the content of the group of quotes that had been assigned to each topic. A list of the most frequent words used within the quotes assigned to each topic was produced, along with a correlation matrix showing the relationships between topics based on the correlation of the maximum *a posteriori* estimates for the topic proportions. STM topics with correlation greater than 0.25 were grouped together as subtopics of a single topic. Topics which were assigned fewer than 5 quotes with theta > 0.3 were not included in the analysis.

The STM process was performed once for the corpus of quotes provided by those who said that they would accept a COVID-19 vaccine, then separately for the corpus provided by those who intended to refuse a vaccine.

### Qualitative Analysis

In each of the two STM analyses, we extracted those quotes that had theta scores above 0.3. This approach was taken on the assumptions that (i) quotes with a high theta score for a specific topic were likely to be highly representative of the topic and (ii) that these quotes were unlikely to have content crossover with other topics because the high theta score was proportional to high exclusivity (noting that the individual topic theta scores for each quote always add to 1.0). Detailed thematic qualitative analysis then allowed us to perform a thorough and nuanced exploration of the topic content and meanings.

## RESULTS

The advert and survey were active between 2020-12-08 and 2020-12-18. The total ‘reach’ of the facebook advert (i.e. the total number of individuals to whom the advert was displayed at least once) included 120,826 people living in England (n = 99,834), Scotland (n = 10,368), Wales (n = 8,384) and Northern Ireland (n = 2,240). The advert reached 69,115 females, 48,534 males and 2,176 people who did not declare their gender on Facebook. The link to the survey webpage was clicked 7,472 times (an overall click-through rate of 0.0618 and cost per click £0.16). In total, 4,535 respondents completed the survey (overall conversion rate from advert to survey completion 0.038, cost per survey response £0.22). There was substantial heterogeneity in the click-through and conversion rates among different age and gender groups.

Demographically, the cohort had notably high levels of representation from females (65%), people aged 50-69 (61%) and people who were educated to degree level or higher (55%). The survey had very low representation of ethnic minority groups (3%) which is in keeping with our findings in a previous (similarly designed) survey [12,18]. Other demographic factors including postcode areas, income and employment were more evenly represented in the cohort (Table 1). The respondents were located in England (n=3,910, 86.2%), Scotland (n=316, 7.0%), Wales (n=263, 5.8%) and Northern Ireland (n=46, 1.0%).

**Table 1.**
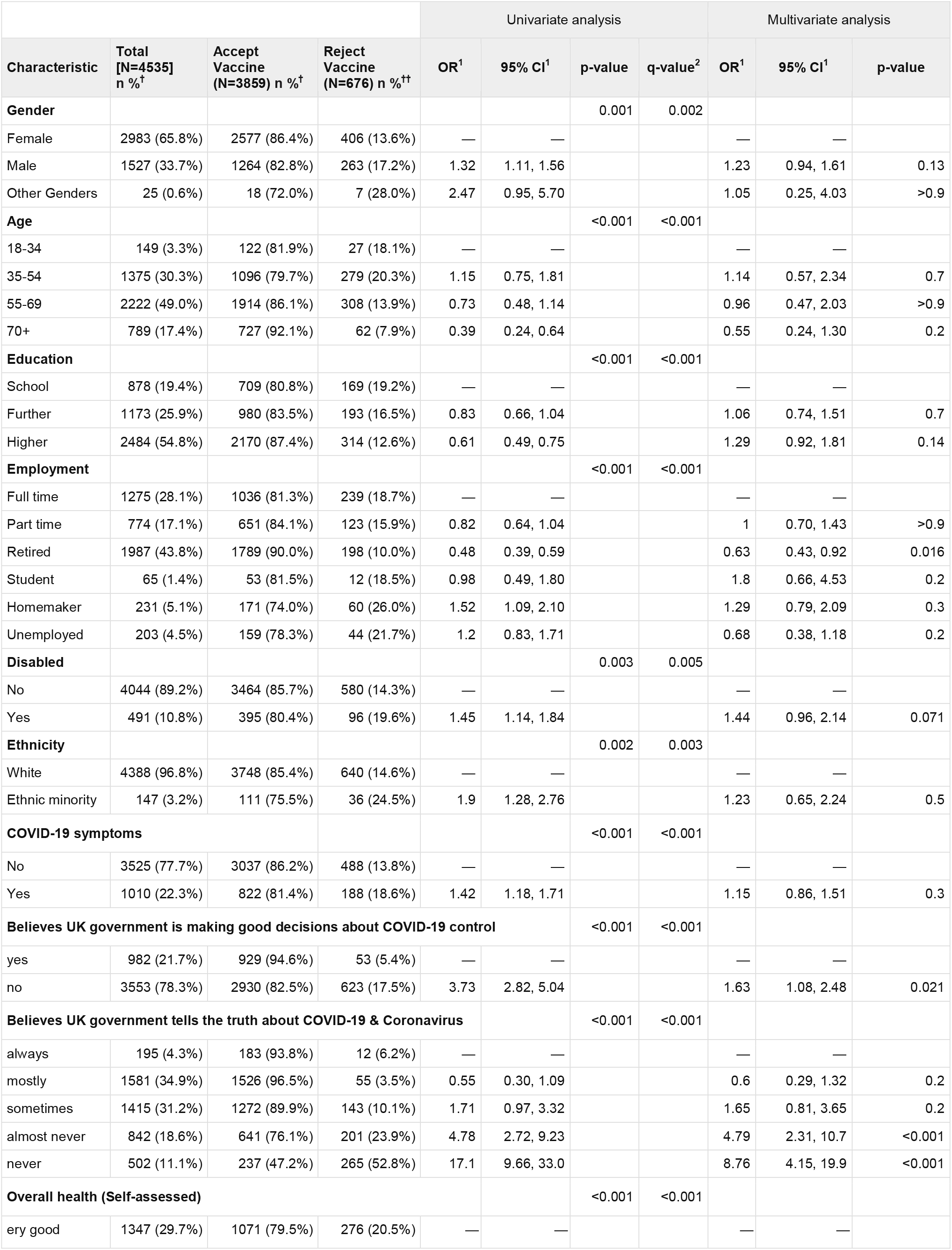
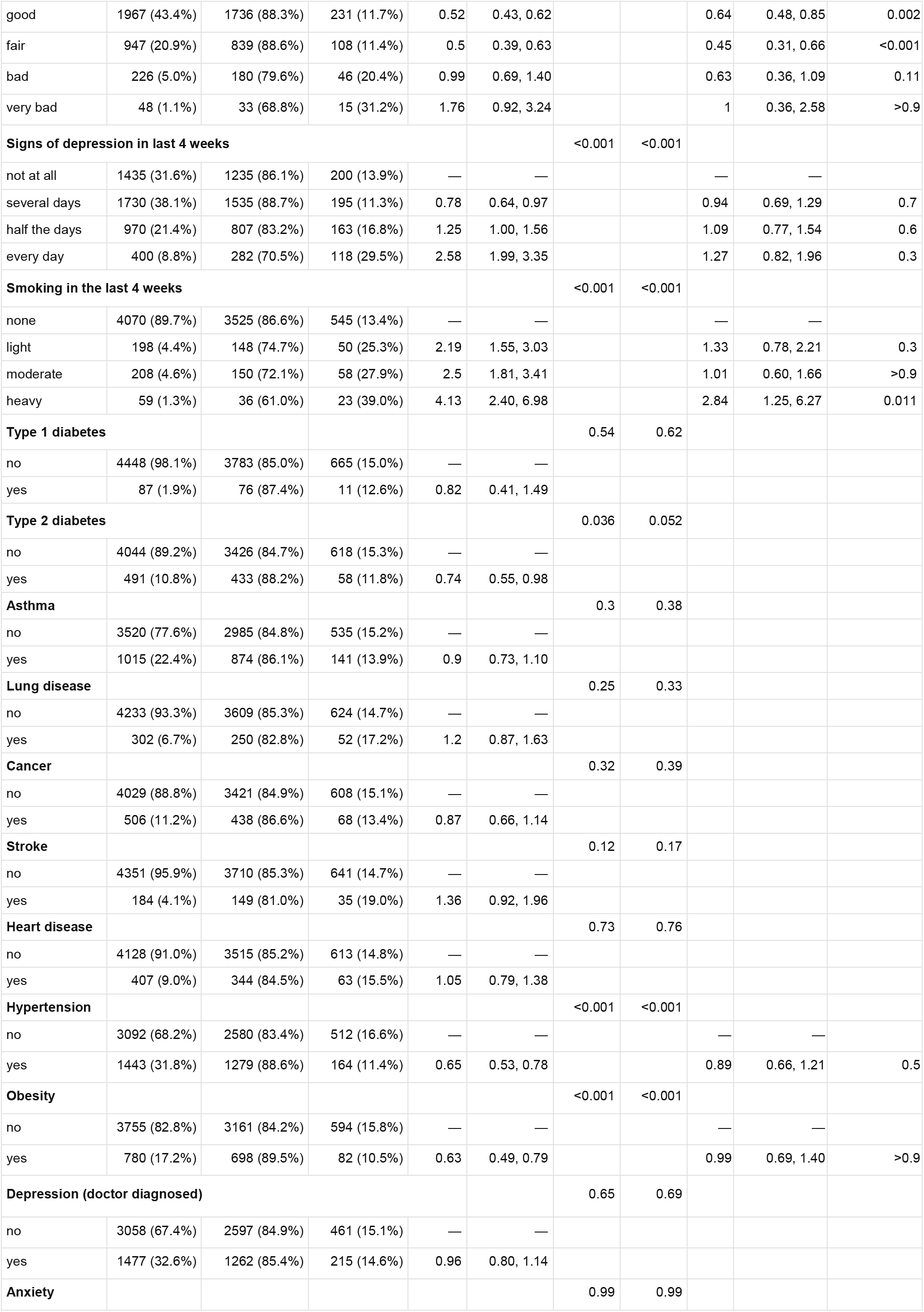
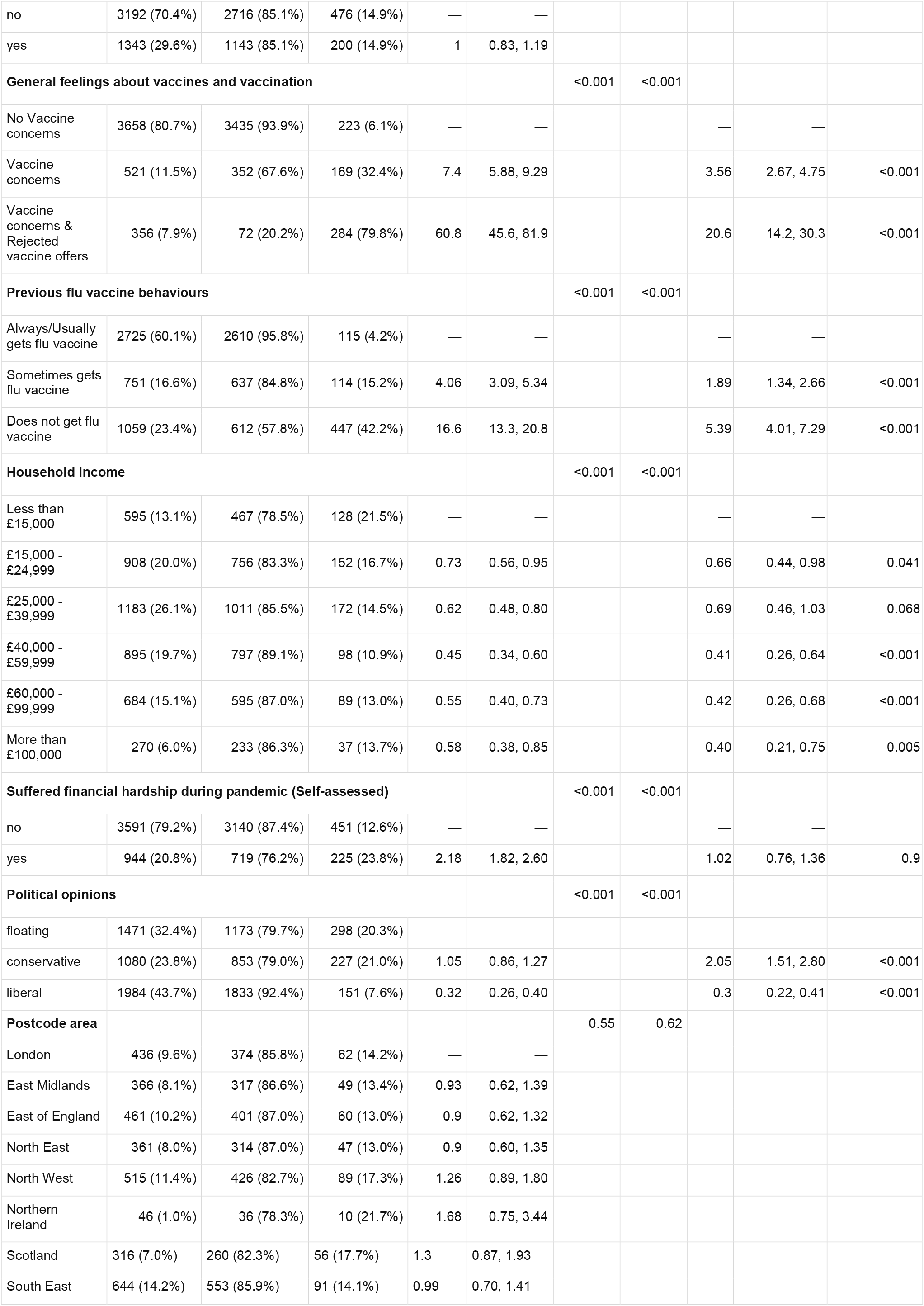
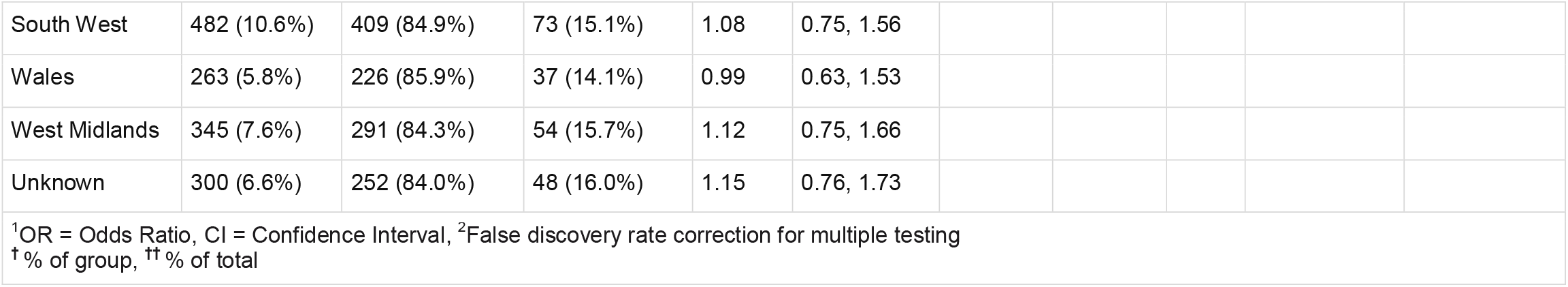
Sample characteristics, univariate & multivariate analyses

Overall, 85% (3859/4535) of participants indicated that they would accept a SARS-CoV-2 vaccine if offered one. Table 1 provides a summary of the survey’s demographic data and the results of both the univariate and multivariate logistic regression tests. Showing only those variables where there were significant differences between the groups in multivariate analysis, Figure 1 illustrates the odds ratios and associated confidence intervals of intending to reject an offer of a COVID-19 vaccine in different groups who took part in the study.

**Figure 1.**
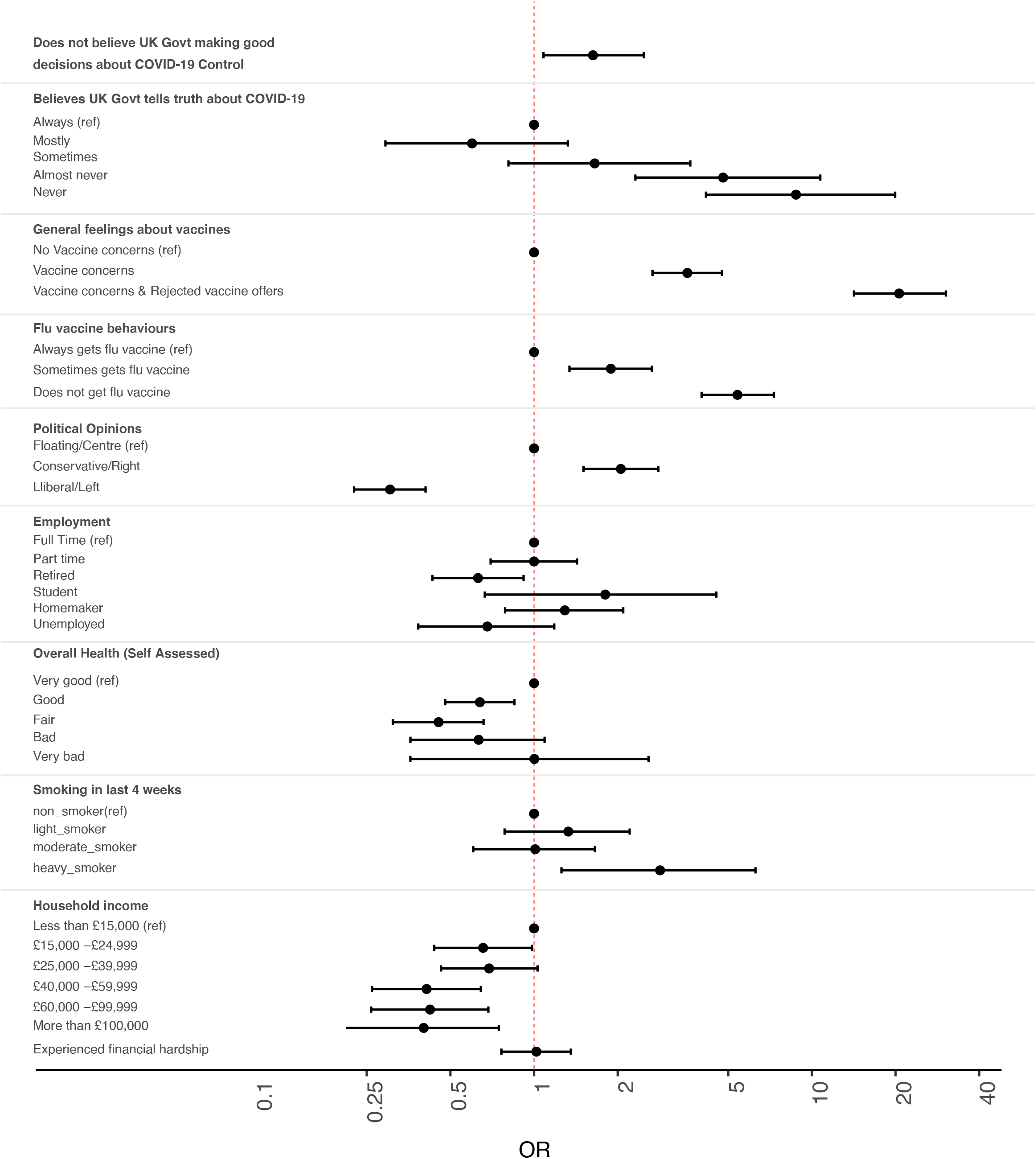
Intention to refuse offers of COVID-19 vaccination among members of the UK public. Odds ratio (OR) estimates [points] and 95% confidence intervals [solid lines] indicate relative intention to refuse vaccine. ORs above 1.00 indicate increased OR of rejecting vaccines. Values below 1.00 indicate increased intention to accept vaccines. All ORs are fully corrected for other covariates. Covariates that had no significant association with intention to vaccinate are not shown, but are fully described in table 1.

The STM analysis identified 16 topics (REF01 - REF16) that together described the key reasons given for refusing a COVID-19 vaccine (Table 2). Topic REF16 had too few quotes and was dropped from the analysis. A further 16 topics (ACC01-ACC16) were identified for the group who intended to accept COVID-19 vaccination (Table 3). Seven sub-topics (ACC02ii, ACC04iii, ACC04v, ACC05i, ACC05ii, ACC05iii and ACC05v) had fewer than 5 quotes with theta > 0.3 and were dropped from the analysis.

**Table 2 :**
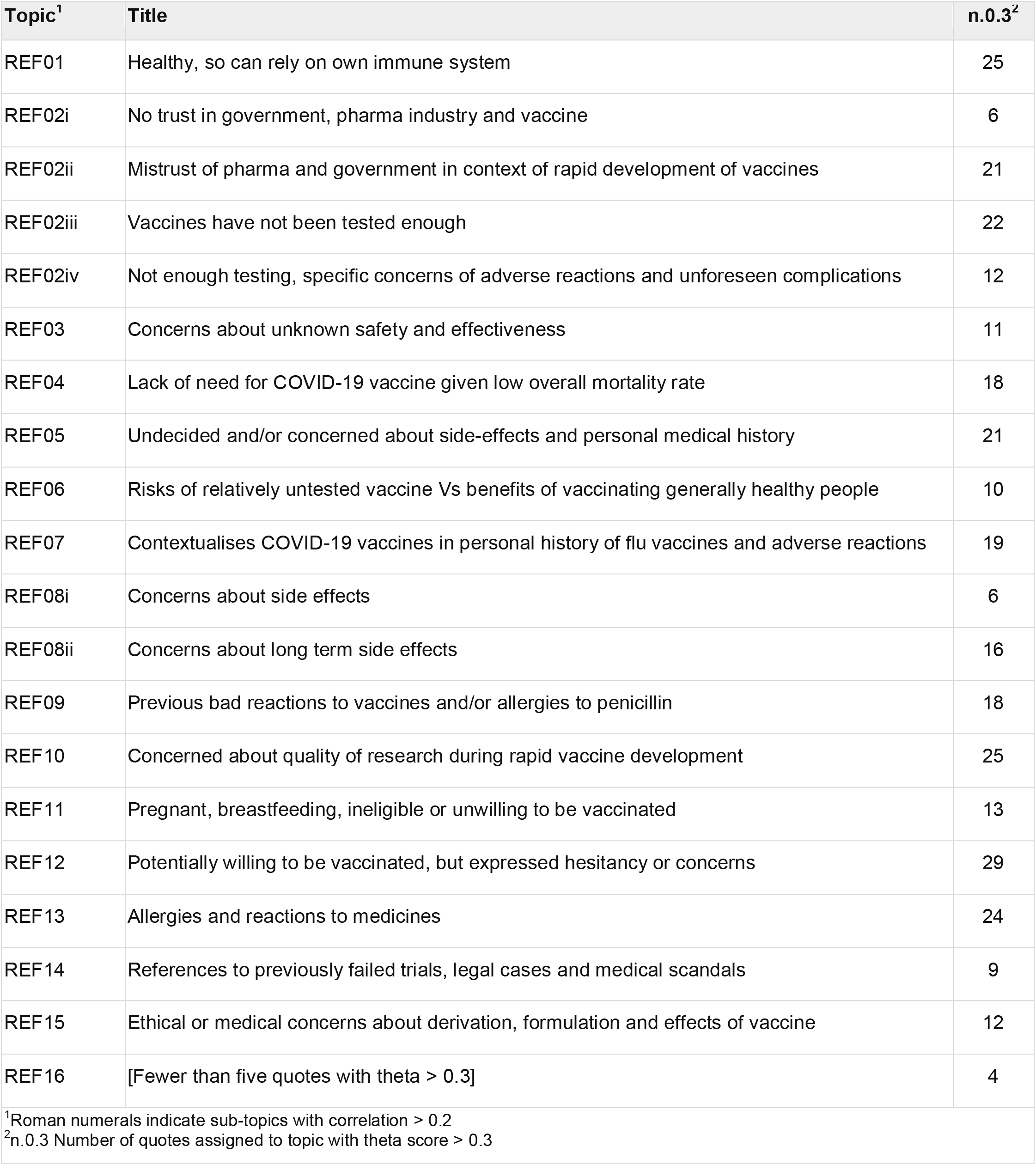
Topics summarising reasons for intending to refuse a COVID-19 vaccine

**Table 3 :** Topics summarising reasons for intending to accept a COVID-19 vaccine

| Topic<sup>1</sup> | Title | n.0.3<sup>2</sup> |
| --- | --- | --- |
| ACC01 | Recognises value of vaccinations and compares COVID-19 to flu vaccines | 37 |
| ACC02i | Wants to get back to normal life | >100 |
| ACC02ii | [Fewer than five quotes with theta > 0.3] | 1 |
| ACC03 | Aware of risks and concerns of others, but on balance sees benefit of COVID-19 vaccines | 39 |
| ACC04i | Wants to protect loved ones / vulnerable people, and travel to see friends and family | 55 |
| ACC04ii | Protect self and others | 7 |
| ACC04iii | [Fewer than five quotes with theta > 0.3] | 1 |
| ACC04iv | Wishes to mix in social, cultural and work contexts | 7 |
| ACC04v | [Fewer than five quotes with theta > 0.3] | 0 |
| ACC05i | [Fewer than five quotes with theta > 0.3] | 0 |
| ACC05ii | [Fewer than five quotes with theta > 0.3] | 0 |
| ACC05iii | [Fewer than five quotes with theta > 0.3] | 2 |
| ACC05iv | To prevent contracting and spreading infection | 8 |
| ACC05v | [Fewer than five quotes with theta > 0.3] | 1 |
| ACC06 | Trusts the science, scientists and the scientific process | 52 |
| ACC07 | Has some concerns, but on balance believes acceptance the correct choice | 13 |
| ACC08 | To help build herd immunity | 58 |
| ACC09 | Recognises vaccination as a social responsibility and public good | 27 |
| ACC10 | Healthcare professionals and/or visit care homes | 20 |
| ACC11 | Responsibilities to the wider community | 17 |
| ACC12 | Works in NHS, Schools, Pharma and with vulnerable people | 34 |
| ACC13 | Participant and/or family member in high risk group because of age or medical condition | 54 |
| ACC14 | Vaccination to be safe and make others safe, some with negative sentiments on UK government | 10 |
| ACC15 | Prevention better than cure, referencing success of historical vaccine programmes | 13 |
| ACC16 | It is the sensible and responsible thing to do | 72 |
| <sup>1</sup>Roman numerals indicate sub-topics with correlation > 0.2<br><sup>2</sup>n.0.3 Number of quotes assigned to topic with theta score > 0.3 |  |  |

### Government decisions

We found that just 21.7% (n = 982) of study participants thought that the UK government was making good decisions about the control of COVID-19. After adjusting for all covariates in a multivariate regression analysis (Figure 1, Table 1), the odds ratios of COVID-19 vaccine refusal were found to be higher amongst those who did not believe that the UK government was making good decisions about the COVID-19 response (OR 1.63, 95% CI 1.08-2.48, p = 0.021).

In topic ACC06, respondents expressed how their intention to be vaccinated was informed by their strong trust in science, the NHS, scientists and scientific method. These respondents were not equally positive about the role of the UK government and some made it clear that although they had doubts about the government’s role, this did not deter them from accepting vaccines.

> *“Although I have little faith in the government, I do have faith in the UK’s medical approval authorities and processes”*
>
> *“I trust science, and our NHS, despite the government’s manipulation of the situation.”*

Among the quotes from potential vaccine refusers, criticisms of the government were generally focussed on questions surrounding their motivations, truthfulness and political or social agenda.

### Government trust

Nearly one third of respondents (29.7%, n = 1,344) believed that the UK government never, or almost never, told the whole truth about coronavirus and COVID-19. The odds ratios of COVID-19 vaccine refusal were higher amongst those who thought that the UK government ‘never’ (OR 8.76, 95% CI 4.15-19.9, p < 0.001) or ‘almost never’ (OR 4.79, 95% CI 2.31-10.73, p < 0.001) told the whole truth about COVID-19.

In topic REF02, participants expressed their scepticism about the UK government’s role in the very rapid process of COVID-19 vaccine development. Many of the respondents whose quotes made up topics REF02 and REF03 felt that it was hard to believe that a valid and legitimate process could be completed in such a short time.

> *“Not enough testing (8 - 12 months compared to 8 - 12 years for normal vaccine development), too much secrecy regarding content and method of production, problems arising even in early cases.”*
>
> *“I religiously get my flu jab each year, and have had numerous other vaccinations throughout my life - the only negative experience was when I was in the NHS in the 1980s when Hep C jabs were being promoted that had been formulated from Us [USA] blood products, but this has not stopped me having all the other proven vaccines above. However re Covid19:- I’m not convinced by the lack of studies of long term effects; need to know more about differences between the vaccines; interested to know how the MHRA had all the info re Pfizer, when the final results were released a week after they had said it was okay; the optics of share sales (moderna)/efficacy results changing within hours of a share price crash (astra zeneca) are not altogether encouraging either.”*

These concerns appeared to reflect suspicions that were implicitly leveled at both the government (REF02) and pharmaceutical companies (REF10).

> *“Because the medical profession tell us that it takes between five and ten years to perfect a good vaccine, we are being told that Pfizer, has done this in six months. Garbage.”*
>
> *“Please! If a cure couldn’t be found for Coronovirus or the other cold viruses since research started in 1947, then ergo, these ‘vaccines’ have obviously already been manufactured well before the ‘pandemic’. My question is Why?”*

Others were suspicious not only of the timeline of development, but also of the potential for personal financial gain for ministers and the alleged provision by the government of legal indemnities to pharmaceutical companies.

> *“No confidence or trust whatsoever in the testing regimes for the vaccines. If they are so good why has [the] government given immunity from prosecution in the event of adverse reactions to big pharma companies?”*
>
> *“Rushed, not properly tested vaccines from companies where there is personal financial gain for govt ministers and friends, also the granting of immunity against liability for adverse drug reactions to all who are developing vaccines, they have been trying to develop a corona virus [sic] vaccine for over 20 years, without success, now suddenly the miracle happens?”*

Some also questioned whether there was any real need to vaccinate the population when COVID-19 was a relatively mild disease in the majority of people and had a low overall mortality rate (REF04).

> *“the virus is mostly a mild disease. Statistics for Covid don’t support general vaccination.”*
>
> *To bring the country to a standstill over a virus with a 98% recovery rate and a death rate one year above the national life expectancy I believe the reaction to it has been grossly over the top I therefore it goes against every sinew in me to get the vaccine I am not scared of the virus and would rather my body battle it naturally and I’m not prepared to have have something that has been rushed through with unknown long-term or short-term side effects put into my body*
>
> *This is a [sic] untested RNA vaccine. Knowone [sic] knows the long terms [sic] effects of this. Plus this virus has a 99.8% survival rate. Therefore vaccines are not needed*
>
> *Too soon to assess effectiveness or side effects. Plus the virus is mostly a mild disease. Statistics for Covid don’t support general vaccination*.

### Confidence in vaccines

The majority of respondents always or usually had a yearly vaccination against influenza virus (n = 2,725, 60.1%). Individuals who reported either never (OR 5.39, 95% CI 4.01-7.29, p < 0.001) or only sometimes (OR 1.89, 95% CI 1.34-2.66, p < 0.001) having a seasonal influenza vaccine were less likely to accept COVID-19 vaccines than those who did so every year. Members of both groups described how they informed their decisions with reference to their use or experiences of flu vaccines. On the side of those who would accept COVID-19 vaccines, topic ACC01 included comments that indicated how some participants saw little difference between the COVID-19 vaccines and those they already used for flu.

> *“I sometimes have the flu jab. Same difference to me”*
>
> *“Same reason I have a flu jab every year - to avoid getting the virus”*

Some participants who planned to get vaccinated (ACC15) reflected on the maxim that “Prevention is better than cure” and cited the successes of previous vaccination campaigns

> *“When I was young polio was common and life threatening. Vaccination changed all that. Presumably this vaccination will do the same for Covid.”*
>
> *“the only disease to be eradicated is small pox [sic] the rest are managed by better treatments and vaccines”*

Meanwhile in the other group (REF07), some participants reflected on their own negative experiences of flu vaccines, focussing on their perceptions of how these had previously harmed their health.

> *“The flu jab caused m.e [myalgic encephalitis] in 2009. I want to build my exposure naturally as when my health improves I will be back at the allotment [a plot of land rented by an individual for growing vegetables]. I understand a small part of virus helps your body fight and remember how to fight in future. I just question what else is in them now. People have glandular fever and are ok. I think it was that and the combination of the vaccine that didn’t give my body a chance. My nanny has also recently said. Bare [sic] in mind she’s 80 she doesn’t feel so well after this recently [sic] flu jab and isn’t recovering…”*
>
> *“I had my 1 and only Flu jab back in 1974 and suffered the worst bout of Flu soon after, 5 weeks off work, over 1 stone lost in weight. I’ve not had a Flu jab in the ensuing 46 years, and I’ve never had the Flu since either, so no, I will not be having the Covid-19 vaccine!”*

The majority of participants (3,658, 80.7%) expressed general confidence in vaccines. Among those who were vaccine hesitant, 521 (11.5%) had concerns about the usefulness of vaccines and a further 356 (7.9%) not only shared these concerns, but had also rejected some or all vaccines they had been offered in the past.

Participants who had concerns about vaccines in general and who either had (OR 20.6, 95% CI 14.20-30.30, p < 0.001) or had not (OR 3.56, 95% CI 2.67-4.75, p < 0.001) previously turned down offers of vaccines against other diseases, were all more likely to indicate intention to refuse COVID-19 vaccines.

Participants who planned to accept COVID-19 vaccines spoke of how they recognised the value of vaccinations (ACC01, ACC05 & ACC14) and of how they accepted that vaccination was a safe and effective process that could help establish herd immunity (ACC08).

> *“I regard it as my personal contribution to the overall health of the population. It is necessary for herd immunity in the true sense of the phrase”*
>
> *“Because herd immunity is essentially a myth: measles, TB, polio, smallpox-none of these (and others) were [n]ever eliminated from a population by herd immunity [presumably meaning by uncontrolled natural spread of infection], but by vaccination.”*

A number of participants made the points (ACC16) that they felt that vaccinating oneself was the “sensible and responsible” thing to do, that (ACC11) these decisions reflected responsibilities to dependents or the wider community in the pursuance of (ACC01 & ACC09) public good through (ACC09) common sense.

> *“Because it’s the right thing to do to see [an] end of how we are all currently constrained”*
>
> *“To protect myself, to reduce the amount of virus in the community, to free myself from isolation”*
>
> *“Because it’s stupid and irresponsible not to unless there’s a genuine medical reason why you can’t.”*
>
> *“We live in a society, each person in that society have [sic] obligations to each other. By having the vaccine you are fulfilling one of those obligations. Each and every person must do what is needed to fight this pandemic no matter what it is otherwise we will never get it under control.”*

Some also stressed (ACC04) that vaccination would protect them and others; including loved ones and social contacts who were vulnerable. For many this was key to returning to normal life (ACC02) and ending social distancing (ACC04).

> *“Protect myself and protect those that can’t be vaccinated. Hopefully enable me to see friends and family again properly and hug them. I haven’t hugged my mum, dad or friends since the start of March and it really hurts.”*
>
> *“I don’t want to get sick and I want my freedom back. I want to see my children and friends, go to the movies or theatre, travel. Have real galleries not online ones. Use public transport etc. without fear.”*
>
> *“it will be the only way to get life back to normal”*
>
> *“I should like to live a normal life. I should like to go on holiday. I should like to sing with other people. I live in London and I’d like to use the Tube, go to the theatre, go to exhibitions.”*

Among the group who planned to accept COVID-19 vaccines, some mentioned that whilst they were not without concerns, they had decided that on balance they could see a net benefit of vaccination (ACC03, ACC07).

> *“I feel I probably should though I am nervous about how ‘young’ the vaccine is. I am aware all stages have been covered in development but it has only been trialed/in use for a very short time. No one has any idea of actual long term side effects. Most vaccines I’d have would have been in use for years. What if we all regret it later? Especially the newer RNA technology. I am more confident in the Oxford style vaccine.”*
>
> *“I don’t think any COVID vaccine will be completely safe as there’s no decades worth of data to assess the long term consequences. However on balance, having the vaccine will be safer than experiencing covid (or passing it on).”*

Others who were willing to be vaccinated explained that they would get vaccinated because they had roles as teachers, healthcare workers or other frontline workers (some with medical vulnerabilities) and/or regularly spent time with people who were vulnerable or in care homes (ACC10 & ACC12).

> *“I work in healthcare and have severe asthma. I don’t want worse lung function or to pass this on to vulnerable relatives and patients. I want tier 3 [social distancing and lockdown] restrictions to end. I don’t want more lockdowns.”*
>
> *“I am vunerable [sic] due to my lung problems. The disease could be fatal for me. I work with very vunerable patients. My mother in law lives with us shes [sic] 85”*
>
> *I am on drugs that suppress my immune system and feel quite vulnerable as I work in school where we can not socially distance from the children”*

Some participants who said they would refuse the offered vaccines clarified that there were in fact circumstances under which they would get vaccinated (REF12). For some, the need to be vaccinated in order to be permitted to travel could be a key influencing factor, whilst others wanted to wait to see longer term effects in others. Still more said that they probably would get vaccinated, but would not be ‘happy’ about it (referencing the specific wording of the survey question, see methods).

> *“I will if I’m coerced by the government to travel but I wouldn’t be happy about it. I’d simply rather not and as I’m low on the list I will wait.”*
>
> *“Has only been tested on [a] small demographic. Company that developed it [has] no vaccine experience. Will happily get London [presumably University of Oxford/Astra-Zeneca] vaccine 2021 after [it is] rolled out to [a] wider group. Plus had Covid recently so [my] assumption is [that I] have antibodies at the moment.”*
>
> *“I think it’s too soon/ quick - has [Prime Minister] boris [Johnson] had it yet? I think it’ll be a while before I get called up so I’ll reconsider then.”*

Several topics (REF02, REF08, REF09, REF10, REF13, REF14) meanwhile triangulated with a general mistrust of vaccines and vaccine research. In topic REF08, participants expressed concerns about the potential for long-term side effects or adverse reactions, especially in the context of the track-records of various pharmaceutical companies.

> *“No evidence that the “vaccine” would actually make a difference at this stage and is unproven in the longer term. I am not statistically at risk and therefore there appears to be more potential issues from the vaccine than from the infection. As more data and evidence emerges over time I will review this.”*
>
> *“Because I don’t feel confident that there’s been an adequate monitoring period after administration. Seeing politicians and the CSO [Chief Scientific Officer] say they would have the vaccine whilst it was still in phase 3 and unapproved destroyed my confidence in it. Pfizer’s track record and the fact they have been granted legal immunity also concerns me.”*
>
> *“untested, unproven, vaccine manufacturers have been given indemnity from prosecution for any harm the vaccine may cause, developed far too fast by companies with a track record of fraud and harm, no long term studies of a brand new vaccine type etc.”*
>
> *“[I am] not convinced at the reassurances by all those involved with their own agendas. Other vaccines have a history of out of court settlements. There is no chance I would touch this vaccine for at least 5-10years.”*

Further to comments indicating a general mistrust of pharmaceutical companies (REF02), some quotes (REF13 & REF14) went on to reference notorious historical medical scandals, including those related to the drugs Thalidomide and Benoxaprofen.

> *“Approval too rushed. I grew up with Thalidomide victims and was myself prescribed Opren [Benoxaprofen], which had to subsequently be withdrawn.”*
>
> *“Until the vaccine has been properly tested* … *I am of the thalidomide generation* … *I would be very wary.”*

Some participants either had personal experiences (REF09) of adverse reactions to medicines or vaccines that had shaped their opinions of COVID-19 vaccines, or were otherwise (REF15) concerned that they could have reactions resulting from pre-existing conditions.

> *“[I am] Slightly concerned and will take advice from GP as have had allergic reaction to flu vaccine and penicillin in the past. More than willing to have vaccine if reassured on likely adverse reaction.”*
>
> *“I had a very bad reaction to a normal flu jab that resulted in going to A&E due to dehydration. The symptoms I had are very similar to the symptoms of covid”*
>
> *“I have Lupus, Hughes Syndrome/APS/Antiphospholipid Syndrome, Sjogrens, a slow thyroid/Hashimotos, Psoriatic Arthropathy and STEVENS JOHNSON Disease, I have dire reactions to drugs, and have an allergy to antibiotics, serious allergies*… *so today we find out that people like me should not have the Pfizer vac, I have had lot of vacs in my life but no more now.”*

Some participants also mentioned their ethical concerns about derivation, formulation and effects of COVID-19 vaccines (REF15, REF08 & REF09)

> *“Vaccines developed using aborted foetal cells are unethical. If there is a vaccine that has been developed without using aborted foetal cells in any part of the manufacture or testing processes, I would be happy to receive that vaccine.”*
>
> *“I react badly to vaccinations. The contents of them are more harmful than the good they might do.”*
>
> *“Will get oxford vaccine when licensed but I have concerns that the Pfizer vaccine utilises muscle to make protein from rna [sic] and that there is no long term back data to show whether this elicits any side effects eg; autoimmune concerns or whether it is totally safe. If I were 80 and high risk I would have it.”*

### Overall Health

Participants who rated their general health as ‘good’ (OR 0.45, 95% CI 0.31-0.66, p = 0.002), “fair” (OR 0.45, 95% CI 0.31-0.66, p<0.001) or “bad” (OR 0.63, 95% CI 0.36-2.58, p = 0.11) were all less likely to refuse COVID-19 vaccines than the healthiest (“very good”) group. These effects were independent of other covariates including age and pre-existing, doctor diagnosed health conditions. In topic REF01, a number of participants who intended to refuse offers of COVID-19 vaccines explained how they felt they could rely on the strength of their healthy immune system or health behaviours to protect them from COVID-19.

> *“Not tested long enough, insufficient test data available, Mnra [sic] vaccine never been licenced before, no trust in the current testing data, i look after myself, i eat well/healthy, exercise, good immune system.”*
>
> *“My diet and lifestyle are good good [sic] and hence immune system is strong”*
>
> *“I have an underlying health condition and vaccinations and medications can compromise my immune system rather than support it. I am not opposed to vaccinations, just cautious and believe in supporting the immune system as much as possible through diet, excercise [sic] and where necessary supplementation ie, vitamin D”*

The concerns of some participants (REF05) reflected a position where they may eventually decide to accept COVID-19 vaccines, but only after further information became available. For some this was guided by their personal medical concerns and overall health, while others who were generally healthy (REF06) found themselves trying to evaluate the risks of accepting a relatively untested vaccine against the benefits of being vaccinated.

> *“[I have] Multiple autoimmune diseases, vaccine not tested on people like me. No evidence that immune response to vaccine won’t cause autoimmune flare or long term immune dysregulation. Not an anti-vaxxer, just a chronic illness sufferer.”*
>
> *“I am undecided, I would like to see more safety data as it is rolled out.”*
>
> *“I would like to see the result of studies into how long antibodies last post vaccination and also learn more about how people are reacting to it. So my answer is NO I won’t take it as soon as it becomes available. By the time my turn comes my answer might well be YES.”*
>
> *“I do not take the flu jab either*… *but I also don’t take any other medication unless it is essential. I do not feel that the long term side effects of things like the flu jab or covid vaccine have been fully investigated or understood. I expect that my usually quite good immune system would handle a coronavirus-if it hasn’t already. It may make sense for people who have been shown to be at higher risk of complications to take the vaccine though. Also, I am pregnant and the impact of the vaccine on fetuses has not been investigated.”*

Others meanwhile considered themselves to be ineligible because they belonged to special demographic categories (REF11 & REF13).

> *“I am breastfeeding. I will get vaccinated if it is safe for my baby or when I stop breastfeeding.”*
>
> *“Trying to get pregnant. I will be happy to get the vaccine once more evidence is provided that it is not harmful to women trying to conceive/pregnant”*
>
> *“I would get vaccinated if I could trust the vaccine and am not an anti vaccer [sic] but I have concerns about the current vaccine in particular the warning to fertile or pregnant women as well as breastfeeding mothers*….. *I am going to fit into one of these categories for many yrs to come!”*
>
> *“I suffer from an ongoing allergy problem and have to carry adrenaline at all times. I understand that this means that I cannot have the Pfizer vaccine. I’ll look at the others when they are released.”*

There was no evidence for a difference in COVID-19 vaccine acceptance among groups with specific medical conditions (as diagnosed by a doctor), those who were disabled, or who had recent signs of depression. Ethnicity was not associated with vaccine intention in this analysis, though we highlight the very small number of respondents from ethnic minority groups and lack of granularity in analysis of this variable. Heavy smokers were also more likely to refuse (OR 2.84, 95% CI 1.25-6.27, p 0.011) offered COVID-19 vaccines.

### Employment status and income

We found that retirees were less likely to decline a COVID-19 vaccine compared to those in full time employment (OR 0.63, 95% CI 0.43-0.92, p = 0.016) but there was no evidence for similar differences in other employment categories. In topic 13, some respondents identified their being older as a key factor in their decision to accept offers of COVID-19 vaccines (ACC13). Others referenced their personal experiences of the impacts of now-vaccine-preventable diseases such as measles (ACC13 & ACC14).

> *“I’m in the older age group and it’s sensible to reduce my risk”*
>
> *“Because of my age and having asthma. Also, my husband has angina, so to help protect him.”*
>
> *“I was born in the 1950s when measles, polio caused severe illness and vaccination changed that”*
>
> *“I still remember when smallpox was finally beaten. I had an older cousin who was blind and brain damaged because her mother caught measles when pregnant.”*

Participants in the lowest household income bracket (< £15,000 per annum) were less likely to accept offers of COVID-19 vaccines than any other income group (Table 1). Experiences of financial hardship during the pandemic were reported by 20.8% (n = 944) of participants, but this was not associated with vaccination intentions (either in the main analysis, or in sub-analyses where [i] income was removed from the model and [ii] where an interaction between income and hardship was included).

### Political views

The political views of respondents were varied, with 43.7% (n=1,984) describing themselves as “Liberal/Left”, 23.8% (n = 1,080) as “Conservative/Right” and 32.4% (n=1,471) as “Floating/Centre” (Table 1). Compared to those whose political views were central, participants on the political left (OR 0.30, 95% CI 0.22-0.41, p < 0.001) were less likely to refuse a vaccine and those on the political right (OR 2.05, 95% CI 1.51-2.80, p < 0.001) were more likely to refuse a COVID-19 vaccine when offered.

## DISCUSSION

The results of this online survey indicate that acceptance of vaccines was, from the very outset of the UK’s COVID-19 vaccination campaign, likely to be very high in all regions of the UK. Around 85% of respondents indicated that they were willing to be vaccinated when the survey took place in December 2020. There appeared to be no relationship between intention to be vaccinated and geographical location, gender, educational achievement, disability, any of a range of specific pre-existing health conditions or experiences of having had COVID-19 symptoms. Participants from wealthier households were more likely to accept vaccines and previous studies have linked lower income to higher levels of uncertainty about COVID-19 vaccines in the UK [19] and elsewhere [20]. An association between reduced uptake of other vaccines and lower incomes has also been seen in other studies in the United States and the UK [21,22].

Other studies in the UK have found that older adults were more likely to accept a COVID-19 vaccine [19,23] but after adjusting for covariates we found no clear association between age and COVID-19 vaccine acceptance. We did however observe that participants who were retired were also more likely to accept vaccination, which may act as a proxy for the same observation about older adults. No other employment status was associated with intention to accept or refuse vaccination.

Despite a known increased risk of severe COVID-19 amongst those with a number of pre-existing medical conditions [24] we did not see any difference in COVID-19 vaccine acceptance between those with and without doctor diagnosed comorbidities. It is therefore possible that the overall perception of health status is more likely to influence respondents’ attitudes towards vaccination rather than their specific medical history. We saw an association between vaccination and self-reported recent general health, where those who felt that their general health was “good” or “fair” were more likely to accept vaccines than those who said “very good”. This fits with observations from the qualitative analysis that some participants felt that they didn’t need to be vaccinated because of their good general health and strength of their immune system. Heavy current smokers indicated that they were more likely to decline a COVID-19 vaccine than current non-smokers. Smoking is linked to a raft of risk taking behaviours, impaired decision making and poor risk evaluations of the future consequences of actions [25] and these considerations could explain the much higher odds of refusing vaccines that we observed among heavy smokers. Mangtani and colleagues [26] recently showed that uptake of influenza vaccines differed by smoking status, whilst Jackson *et al*. [27] highlighted that current smokers were more likely to be undecided or unwilling to receive a COVID-19 vaccine. The latter study also showed that smokers were more likely to have negative feelings towards vaccines, citing reasons that broadly echoed the concerns raised by the group of participants who planned to refuse vaccines in this study.

The diminishment of public trust in the UK government has been a major concern during the COVID-19 pandemic and we have previously highlighted how generalised mistrust, concerns about the transparent use and communication of evidence and insights into decision-making processes affected perceptions of the UK government’s early pandemic response [28]. In the April 2020 survey, we found that 42.3% of study participants thought that the government always or nearly always told the truth about COVID-19 and in this survey this remained relatively unchanged at 39.2%. Confidence in the quality of government decision making had however changed substantially between the two surveys. In April 2020, 52.7% of participants said that the government was making good decisions [28], but in December 2020 just 21.7% felt this way. A poor opinion of government decision making (and also low trust in the truthfulness of government) was associated with increased odds of refusing COVID-19 vaccines (Table 1). Some respondents who talked about the government in the context of trust did however make a clear delineation between their faith in the government and that in the health service and/or science and scientists.

We also observed that participants who identified as having right-wing political views were significantly less likely to accept a COVID-19 vaccine, even after adjusting for potential socio-demographic covariates. The opposite was true of participants from the political left. Surveys conducted in the US also found that those whose political view was conservative/right were less likely to accept vaccines (28,29). This finding may be somewhat surprising because the UK’s COVID-19 response was led by a Conservative government and we might have expected that supporters of the incumbent government would most positively support their health initiatives. Anti-vaccination sentiments have however been linked with support for populist political parties, with both phenomena being “driven by similar dynamics: a profound distrust in elites and experts” [29]. The rise of populist narratives, and particularly the UK government’s courtship of a populist right wing during the Brexit period (2016-2021) may therefore have led to the consolidation of political but not policy support amongst certain sections of the Conservative electorate.

Previous behaviours and experiences with regards to utilisation of vaccines against seasonal influenza were associated with increased uptake of COVID-19 vaccines. This finding has also been seen in other studies in the UK [19,23], Australia [30] and (in some demographic groups) in the United States of America. [31]. Perhaps unsurprisingly, our results also identified that participants’ levels of generalised vaccine hesitancy and previous refusals of offered vaccines associated with respectively 3.6 and 21 times greater odds of refusing COVID-19 vaccines, compared to those who were confident in vaccines.

During the pandemic, the significance of so-called “infodemics” and of misinformation, rumour and unscientific beliefs, has been at the forefront of public debates [32]. Underlying these discussions is the assumption that hesitation towards or refusal of public health interventions is grounded in misconceptions or poor information. We found only scant evidence within our corpus of free text quotes to support concerns that intentions to refuse COVID-19 vaccines were potentially driven by effects relating to misinformation or pseudoscience. Instead, a key element in our respondents’ considerations was trust in the product, with responses amongst both those who said they would and those who say they would not take a COVID-19 vaccine gravitating around questions about the quality and safety of the vaccines. This supports critical insights into vaccine hesitancy which reject “knowledge deficit” framings of the problem, recasting it instead into a question of trust in scientific expertise [33].

Among those participants who said they would not accept a COVID-19 vaccine, we found that their primary concerns were highly specific to the vaccine development process, to issues relating to the rapidity of the vaccine trials & the novelty of mRNA based vaccines; as well as to the relative absence of knowledge about the long-term effects and safety including in pregnancy and breastfeeding. Whilst the seemingly low penetration of misinformation in our corpus of text may have been biased downwards by the limited representativeness of our sample, empirical social scientific research on rumours, hesitancy and trust in medical research across different contexts has shown similar results [34]. In the context of the concerns of this study’s participants, COVID-19 vaccine hesitancy appeared to be framed by legitimate concerns about the perceived unavailability of substantive scientific data to support claims that the vaccines were safe and (possibly to a lesser degree) effective. It is notable that one COVID-19 vaccine had received licensure in the UK at the time of the survey and that specifically long-term safety issues were a focus point of public discussion.

The safety of vaccines is a prevalent topic in the debate on vaccine hesitancy [35–37]. Reduced confidence may be heightened when limited scientific data are available, for instance in the case of investigational, experimental, and new vaccines used during the response to epidemics [38]. Some respondents claimed that whilst they would not accept a vaccine at present, they might possibly do so in the future, for example when there was a better understanding of potential adverse events following vaccination in other members of the population. The timing of data collection is salient here, as our respondents’ reflections show the particular considerations surrounding deployment of a new vaccine during a health emergency. Indeed, this desire to defer vaccination reflects similar observations made during studies on the acceptance of a vaccine against Ebola virus in Sierra Leone [39]. Some governments even made decisions to defer COVID-19 vaccination at the level of national policy [40]; for instance by delaying the commencement of COVID-19 vaccination programmes whilst observing the impacts of programmes underway in countries with a higher burden of disease [40].

If concerns around vaccination were very specifically related to questions about vaccine development, then reflections on willingness to take the vaccine amongst our respondents were rooted in expressions of social responsibility and a vision of the public good. Many respondents focused on the desire to return to ‘normality’, to be able to see family and friends, to resume social activities and to be able to travel again. These motives for willingness to be vaccinated with a novel vaccine represent a complex interaction between altruistic actions and self-interest that have been reported by others participating in experimental studies [41,42].

Key limitations of this study were that it was non-representative and that the sampling-method was not random; meaning that the study findings are not generalisable either in the UK or elsewhere. We caution in particular that whilst vaccine dis- or mis-information did not appear to be a major influencer of vaccine choice in this UK cohort, this may not be the case in other jurisdictions. The study’s participants were disproportionately likely to be highly educated, white and aged over 50 years. Ethnic minority groups are at greater risk from COVID-19 [24,43] and are also less likely to report that they will accept COVID-19 vaccination [44] but these groups were under-represented among those responding to the survey. Facebook’s advertising policies preclude the targeting of boosted posts to specific ethnic minority groups, and we were therefore unable to influence the degree to which the advert was seen by, or engaged with by members of ethnic minority groups. The study was observational and causal links between the outcomes and statistically associated explanatory variables cannot be assumed. Additionally, covariates which were not included in the study, or any that were misclassified, could have led to residual confounding. As the study relied on self-reported information, there was scope for response bias, although we designed the questions to minimise this wherever possible. Finally, whilst the STM analysis is fully reproducible using statistical software, the assignment of topic names was performed manually; albeit only after a process of discussion and consensus that included all authors of this work.

## CONCLUSIONS

Our embedded mixed-methods analysis highlights groups that may be less engaged with the vaccination program and also provides a more detailed exploration of the complex factors and considerations that influenced decision-making among those who planned to either accept or reject the offered vaccines.

We suggest that in the context of the COVID-19 pandemic, levels of public complacency about SARS-CoV-2 vaccines were very low; whilst the large scale, well organised and equitable NHS-led deployment of vaccines in the UK meant that convenience was high. The majority of respondents to this survey reported on how they intended to accept offers of vaccination, suggesting that confidence was high, regardless of the many unknowns. The dominant theme among all topics relating to vaccine acceptance was that there was a need to restore normality, and for individuals to participate in the social good of vaccination by protecting themselves, their loved ones and others in their communities.

Reasons for refusing offered vaccines were more diverse, but notably focussed on legitimate questions about the safety and efficacy of vaccines, the speed of their development cycle, whether there was a real need for a vaccination program and to a lesser extent on whether the government and pharmaceutical companies had hidden motivations. There was no evidence of widespread misinformation or factors relating to an infodemic having influenced the decisions of many participants.

Engaging with public perceptions of a newly developed vaccine, deployed at the height of a health emergency, we question whether the dominant narratives of vaccine hesitancy are misconceived as primarily (or dominantly) being driven by ‘fake news’, misinformation and disinformation on an epidemic scale (infodemics). Whilst we fully recognise the importance of combating misinformation, we propose that efforts to maintain high levels of vaccine confidence should not neglect to contend with legitimate concerns about the lack of transparency about the scientific process of vaccine development. Public engagement should therefore attend to deeper questions of (mis)trust in science and leadership. In order to maximize uptake, the vaccination programme should leverage the sense of altruism, citizenship and collective responsibility that motivated many of our participants to get vaccinated.

It should be noted that this study was conducted in December 2020, at the start of the mass vaccination campaign in the UK. Current attitudes to vaccination may have subsequently changed, but the aim of this paper is to contend with perceptions of a novel vaccine being prepared for deployment at the height of a health emergency. We reiterate the call of Scheinerman and McCoy (2021) to address these issues through effective engagement with the public through a process of transparency, ethical reasoning and both formal & informal deliberation [45].

## Data Availability

All data and scripts required to reproduce analysis will be available through LSHTM Data Compass (https://datacompass.lshtm.ac.uk/) after institutional review of our submission (Item 2337)

## ETHICAL STATEMENT

This study was approved by the research ethics committees of the London School of Hygiene & Tropical Medicine (ref: 17860) and World Health Organization (ref: CERC.0039B). The data were fully anonymous and the study team were unable to identify any respondents. The respondents provided informed consent at the start of the survey by means of ticking a box on the web-form. All questions in the survey were optional, meaning that participants could skip questions if they did not want to divulge specific data.

This work received funding from the World Health Organization COVID-19 R&D Blueprint Roadmap. Grant number CERC.0039.

## PATIENT AND PUBLIC INVOLVEMENT

A website with the findings was made available to the public. The survey methods were guided by ongoing work with the open-source software development community which maintains and develops the ODK project.

## AUTHOR CONTRIBUTIONS

ChR, HB, LE & SL conceptualized the study. All authors designed the survey. ChR curated the data. ChR, HB, LE & SL performed the analysis and wrote the manuscript. All authors contributed to the article and approved the submitted version.

## ACKNOWLEDGEMENTS

The study team would like to thank the study participants for their contributions to the data and to acknowledge the invaluable administrative support of Eleanor Martins and Rosa Arques.

## DATA AVAILABILITY

The datasets and analysis scripts used in this study can be found in online repositories. The names of the repository/repositories and accession number(s) can be found below

LSHTM Data Compass (datacompass.lshtm.ac.uk) : Item 2337 [DOI Pending]

